# Associations and mechanisms of influence between climate variables and norovirus seasonal incidence: a systematic review and meta-analysis

**DOI:** 10.64898/2026.02.26.26347190

**Authors:** Igor Pântea, Andrew J. K. Conlan, Katy A. M. Gaythorpe

## Abstract

Incidence of norovirus has strong seasonality in temperate and continental climates. Many studies have examined its association with climate variables, but evidence remains disparate. We address this gap by performing a systematic review to summarise and interpret the strength and directionality of associations between climate variables and norovirus incidence.

Embase, Scopus, Web of Science and PubMed databases were screened for peer-reviewed studies on 2^nd^ of December 2024. Articles were included if they described any climate or meteorological variable, in a categorical or numerical format, relative to a measurement of norovirus incidence risk in a human population, or prevalence or survivability outside the human host. Bias was assessed using a modified Critical Appraisal Skills Programme checklist. If dispersion of the effect in a human population was provided, the mean size was calculated using inverse variance weighting. The effect size outside the host was summarised as D-values, representing the time required to achieve a 90% reduction in the detected amount of virus.

A total 139 studies were included. Predictors of risk were ambient and water temperature, relative and absolute humidity, anomalies of ambient temperature and precipitation, atmospheric and vapour pressure. High heterogeneity in direction and size of effects was observed due to regional differences in the factors driving norovirus seasonality and differences in outcome and exposure definitions. Our review suggests that the sensitivity of norovirus to individual climate variables is region and time specific, reflecting geographical differences in the relative importance of norovirus transmission via environmental pathways versus human-to-human contact.

**Plain Language Summary:** Norovirus, a gastrointestinal virus, has a higher number of cases during specific months of the year. Regions with similar types of climate appear to have similar time periods when the increase in the number of infections occurs, which has been linked to norovirus case numbers being correlated to individual climate variables, such as temperature or rainfall. To understand how these associations compare globally and what are their potential explanations, we screened four major scientific databases, namely Embase, Scopus, Web of Science and PubMed. After the selection process, a total 139 peer-reviewed studies were included in this study. We found that ambient and water temperature, relative and absolute humidity, anomalies of ambient temperature and precipitation, atmospheric and vapour pressure were predictors of an increase in norovirus cases. However, the strength and direction of the relationships differed from region to region. A potential explanation is that geographies also differ in how important individual routes are for the transmission of norovirus, specifically via the environment as opposed to direct human-to-human contact, whereas climate is likely to have a greater influence on the former.

**Key points:**

1. The strength and direction of associations between climate variables and norovirus incidence varies by region and time period
2. The strength of associations vary across the transmission routes of norovirus, e.g., environmental versus human-to-human contact
3. Climate variables impact norovirus survival and dissemination outside the host, which may inform models of environmental virus transmission

## 1. Introduction

Norovirus (NoV), a gastrointestinal pathogen also referred to as the winter vomiting virus, is responsible for 18% (95% confidence interval (CI): 17-20%) of all cases of diarrheal disease globally (Lopman, 2015). Annually, it is estimated to cause approximately 685 million acute cases of gastroenteritis and a global economic burden of $64.5 billion (Bartsch et al., 2016; CDC, 2024). NoV burden is seasonal in many countries, and specific months of the year have higher case incidence than other months; hence, by being able to project seasonal trends, healthcare services can implement appropriate preparation measures. However, the predictors and the drivers of NoV case seasonality remain uncertain.

Studies have attempted to link seasonality of NoV incidence to seasonal climate variables, but it is unclear whether the climatic sensitivity of NoV incidence trends is consistent among different regions. Based on results published between 1997 and 2011, Ahmed, Lopman & Levy (2013) found that 78.9% of NoV cases occurred in the colder periods of the year, that is October to March in the Northern Hemisphere and April to September in the Southern Hemisphere. A significant limitation is that studies that associate colder months with higher NoV burden are more likely to be in regions with temperate and continental climates (Ahmed, Lopman & Levy, 2013; Mounts et al., 2000). In dry and tropical climates, such as Sub-Saharan Africa, seasonal patterns of NoV incidence are inconclusive with variations between regions and years, and NoV surveillance data is lacking, especially for adults and older children (>5 years old) (Munjita, 2015; Lopman, 2015; Mans, 2019). A second limitation is that cross-regional comparisons of NoV incidence seasonality are made only relative to ambient temperature or precipitation, whereas relationships or cofounding with other climate variables remain neglected (Chua et al., 2022). A third limitation is that previous comparisons included primary research only with identical measurements of outcome, i.e., relative risk, whilst excluding evidence from studies with other types of effect. Therefore, in this review, we summarised evidence from primary research that described the sensitivity of NoV incidence to individual climate variables with the aim of uncovering if the relationships are consistent across regions with different climate profiles. Firstly, if these associations are consistent in regard to directionality this would provide more support for climate variables driving the seasonality of NoV rather than climate being only a temporal predictor with no mechanistic influence (Levy et al., 2016). Secondly, understanding whether NoV sensitivity to climate is a global phenomenon can facilitate assessments of climate change, such as increased winter temperatures, changes in rainfall patterns, number of floods, or an increased number of extreme weather events (Rohayem, 2009; Patz et al., 2003; Campbell-Lendrum, Corvalán & Prüss– Ustün, 2003).

We also systematically reviewed environmental and laboratory studies that described the relationship between climate variables and NoV detection outside the host. Since, in addition to direct person-to-person transmission, NoV transmits by environmental routes, understanding the impact of climate variables on NoV presence and viability in the environment could aid the understanding of the occurrence of disease in human populations.

## 2. Materials and Methods

### 2.1. Search, Selection and Extraction Strategy

The protocol for screening, including inclusion exclusion criteria, was published on PROSPERO with the ID CRD42024628722 (Pantea & Gaythorpe, 2024). The review followed PRISMA 2020 guidelines to ensure risk of bias and sensitivity analysis are completed and reported appropriately. Briefly, databases Embase, Scopus, Web of Science and PubMed, which includes Medline, were searched on the 2^nd^ of December 2024. The search strategy was refined to a broader terminology so as to identify and include all meteorological or climate related papers, that is *“(norovir* or norwalk*) and (environment* or climat*)”* with no date restrictions. Grey literature, such as government reports or academic dissertations, was searched for using Google Scholar and the OpenGrey database, but none qualified for inclusion.

Two reviewers were involved in the study design and screening process. To evaluate the similarity in their interpretation of the inclusion and exclusion strategy and the aims of this systematic review, a Cohen’s kappa test (Eq. 1) was performed on 400 articles (9.32%) of the total 4,294 found deduplicated articles. It accounts for probability of agreement occurring by chance, where:

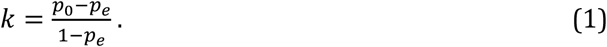

In the equation, *k* = Cohen’s kappa coefficient, *p*_0_ = relative agreement between reviewers, and *p*_*e*_ = probability of agreement between reviewers by chance.

The Collaboration for Environmental Evidence guidelines for systematic reviews recommends a threshold of 0.5 before starting the screening (Collaboration for Environmental Evidence, 2013). After confirming sufficiently good agreement, two reviewers, independently of each other, screened all the articles using titles and abstracts against the inclusion and exclusion criteria, after which the included articles were screened for full text. Conflicts at any stage were resolved by reaching consensus. Extraction of data from the final included studies was split among the authors and was performed according to a standardised form.

For inclusion, we considered any meteorological or climate variable with at least two qualitative or quantitative datapoints described in relation to a measurement of NoV. Epidemiological and environmental studies were included, as well as laboratory studies with climate-like exposure conditions, such as temperature, relative humidity, or ultraviolet radiation. For laboratory studies, experiments may have used surrogates of NoV due to the difficulty to culture human NoV in vitro. In the final dataset, the included surrogates of human NoV were murine NoV (MNV), feline calicivirus (FCV), canine calicivirus (CaCV), and F-specific or male-specific bacteriophage and its subtype MS2 due to only these being described in the context of human NoV in the dataset of the deduplicated studies. For more details on the inclusion and extraction strategy, please refer to the protocol on PROSPERO (Pantea & Gaythorpe, 2024).

### 2.2. Risk of bias assessment

Each individual included study was evaluated using an adapted Critical Appraisal Skills Programme (CASP) assessment (CASP, 2025) in order to identify if the quality and risk of bias of that study requires its results to be interpreted cautiously or to be excluded from the meta-analysis. The categories were “Good”, “Adequate”, or “Poor” across nine criteria relevant to the aims of this systematic review. The assessment was of the 1. study aims, 2. appropriateness of methodology, 3. appropriateness of methodology for the stated aims, 4. number of replicates or measurements to minimise the play of chance, 5. presentation of the main result, 6. procedure of data collection in order to address the research issue, 7. rigour of data analysis, 8. clarity in stating the findings, 9. value, accuracy, and importance of research.

For a final “Good” score, studies required no “Poor” qualifications and a maximum of two “Adequate” results. For an “Adequate” qualification, no “Poor” and a minimum of three “Adequate” scores were required. For a final “Poor” classification, studies required a minimum of one “Poor” result in any of the nine categories.

### 2.3. Meta-analysis

To be able to visually compare the results across different studies and metrics, the measurements of NoV burden were converted to common units where possible. For epidemiological studies, measurements of risk, specifically relative risk, rate ratio, regression slope, and prevalence ratio, were converted to percentage change. Effects were either dichotomised or continuous, where the first represents percentage change of NoV risk in the population between two groups of an individual climate variable, and the latter represents percentage change for an increase in one unit of predictor variable for that type of metric. However, for performing the meta-analysis and summarising the mean effects, the original mean effect sizes defined by the authors of the study were used. No conversions were conducted for environmental studies as these were used narratively to interpret associations of climate variables to NoV incidence in the human population. For laboratory studies that assessed virus reduction, the common units were D-value, which represents the time (or intensity in the case of ultraviolet light (UV) light) required to achieve a one log10, or 90%, reduction in the detected amount of virus. Articles may have used an alternative terminology, e.g., T90 or D10-value, which was extracted as a D-value. When not provided, the D-values and the corresponding 95% CI were calculated, as described in Supplementary Materials 1 “2. Calculation of D-value”, from other metrics of virus concentration.

For summarising the effect sizes of the association between individual climate variables and measurements of NoV incidence in the human population, we used a weighting methodology based on the inverse of sample variances. The procedure described previously (Tang, Caudy & Taxman, 2013; Sánchez-Meca & Marín-Martínez, 2008) is started by obtaining the mean variance of the mean effect size from individual studies:

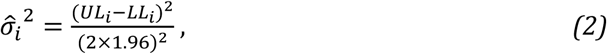

where 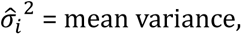 *UL*_*i*_ = upper 95% CI, *LL*_*i*_ = lower 95% CI, *i* = the study index, and the distribution of the mean effect size is assumed to be normal. Equation 2 can then be followed by calculating the mean effect size with:

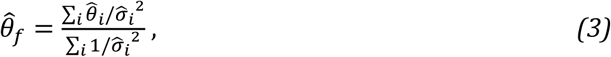

where 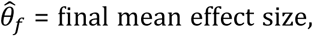 and 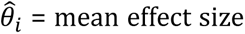of the study with index *i*. The mean variance is then obtained from:

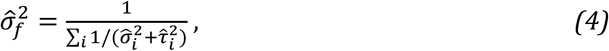

where 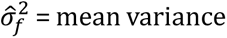of the final mean effect size, and 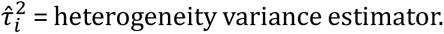 Studies generally did not report 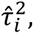 hence, throughout this systematic review, it was assumed to be zero.

Some studies, that used a regression slope coefficient as the measure of effect, did not provide measures of standard error, standard deviation, variance or CI for the mean effect size, hence were excluded from the meta-analysis. Inverse variance weighting was not applied for summarising coefficients of pairwise correlations; in which case, a simple average formula was used.

The meta-analysis for the human populations was run initially for studies qualified in the risk of bias assessment as “Good” and “Adequate”. The presented results also excluded the relationships marked as “insignificant” and including those marked with “significant” and “significance not tested”. For sensitivity analysis, the studies qualified as “Poor” were included too, and an additional analysis was performed on studies qualified as “Good” and “Adequate” but only of a “significant” level. No funnel plot was performed due to heterogeneity in the types of the effects and definitions of individual climate variables.

For systematising and analysing the evidence from laboratory studies regarding the impact of climate-like exposures on NoV survivability and reduction rates in the number of infectious units outside the host, we grouped the evidence by epidemiologically relevant mediums, specifically, fomites, liquid solutions, aerosols, faeces, manure and slurry, and food supply chain. Data was displayed as box plots with average D-values provided narratively. The box plots and average D-values were obtained by type of assay, that is infectivity or molecular assays, where the first involved a preliminary step of culturing viruses that are viable and the second implied virus quantification without culturing it in a cell-line (Supplementary Materials 1, “1. Infectivity and molecular assays”).

All data analysis was performed with R software (version 4.3) with the code and an additional copy of the extraction file (Supplementary Materials 3) being available at github.com/mrc-ide/climate_norovirus_litreview.

## 3. Results

### 3.1. Overview

The level of agreement between the two reviewers for the inclusion and exclusion criteria based on Cohen’s kappa was 0.65, confirming substantial agreement. The PRISMA flowchart is shown in Figure 1. A total of 8,690 potential studies were identified through database screening and literature searching. After removing 4,396 duplicates, 4,294 articles were screened using titles and abstracts, among which 244 were screened for full text. A final dataset of 139 articles (Table S7) was included in the review after removing 105 studies due to the environmental variable being not relevant, the association between the measurement of NoV and the environmental variable not being analysed or not applicable to the aims of this review, the study presenting no new primary research results (e.g., systematic reviews) or not having been peer-reviewed (e.g., conference abstract), the impossibility to access the full-text, or the full-text not having been able to be interpreted from another language despite the abstract being in English. The extraction file containing each study’s characteristics, risk of bias, direction and significance of the association, numerical or categorical boundaries of the exposure variable, an effect estimate where available, a mechanism of influence, and other attributes of the association between climate variables and risk of NoV in the population or in an environment outside the human host can be found in Supplementary Materials 3 (Table S7-S17).

**Figure 1.**
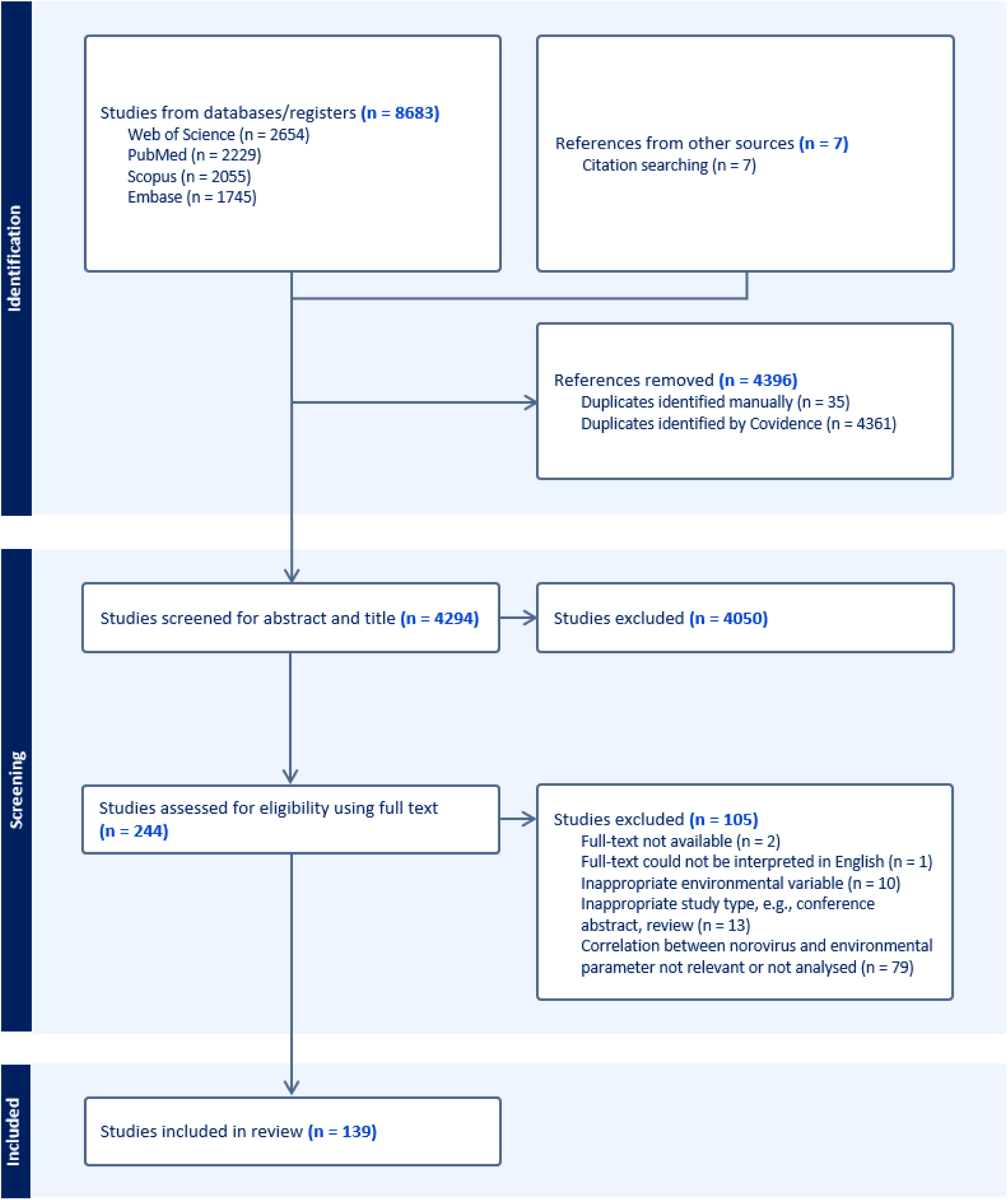
PRISMA flowchart: number of studies extracted and removed during screening against the inclusion and exclusion criteria.

The earliest study that was included was published in 1999 (Figure 2) with environmental and epidemiological studies being distributed across all continents but with no studies in Northern and Central Africa, Western Asia, and limited evidence for Central, Eastern and Southeastern Europe, and Southeast Asia (Figure 3; Table S1). The geographical distribution in Figure 3 excludes laboratory studies and mesocosm-based environmental studies that were not exposed to natural “outside” conditions.

**Figure 2.**
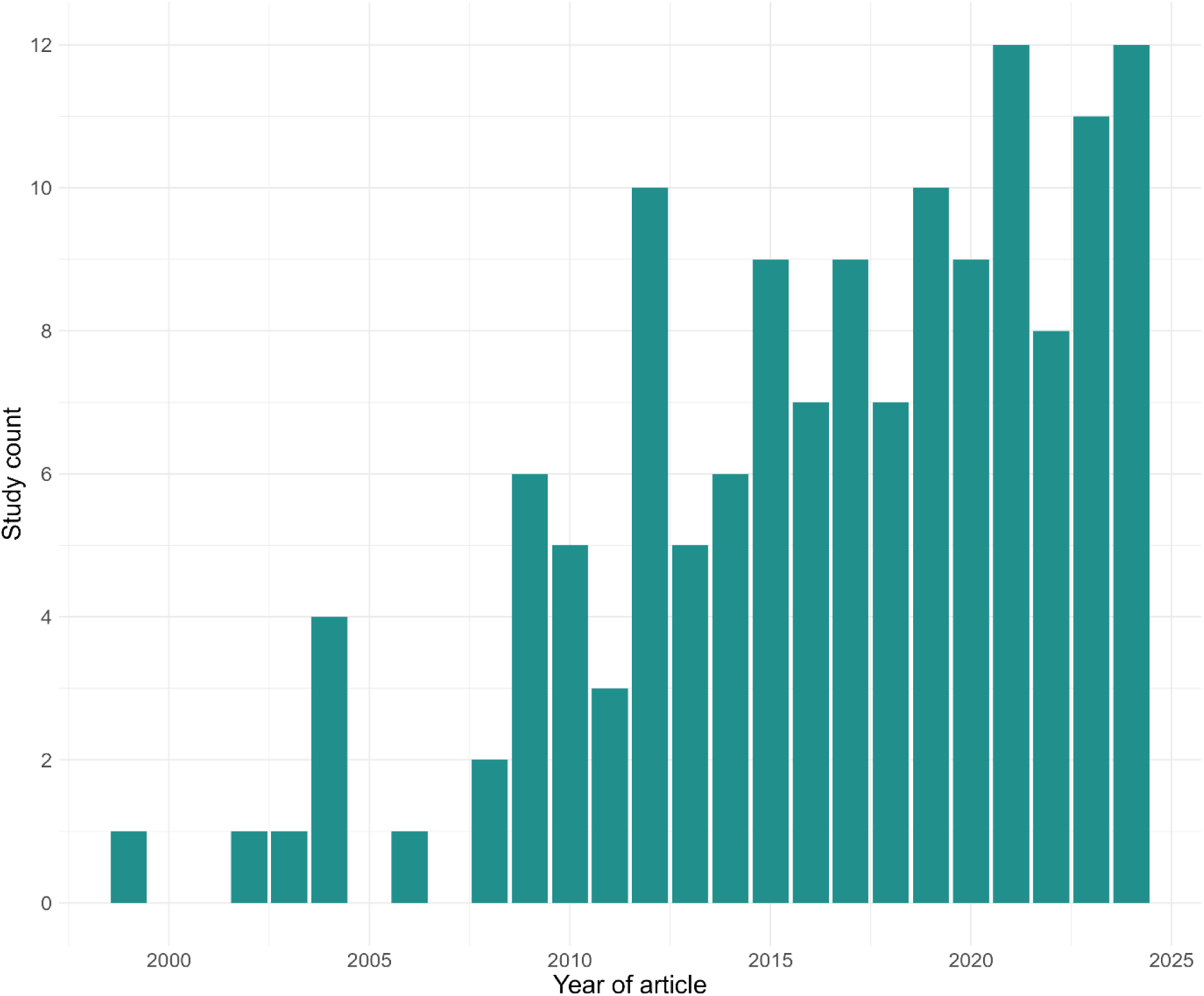
Study count by year of publication of the included 139 studies.

**Figure 3.**
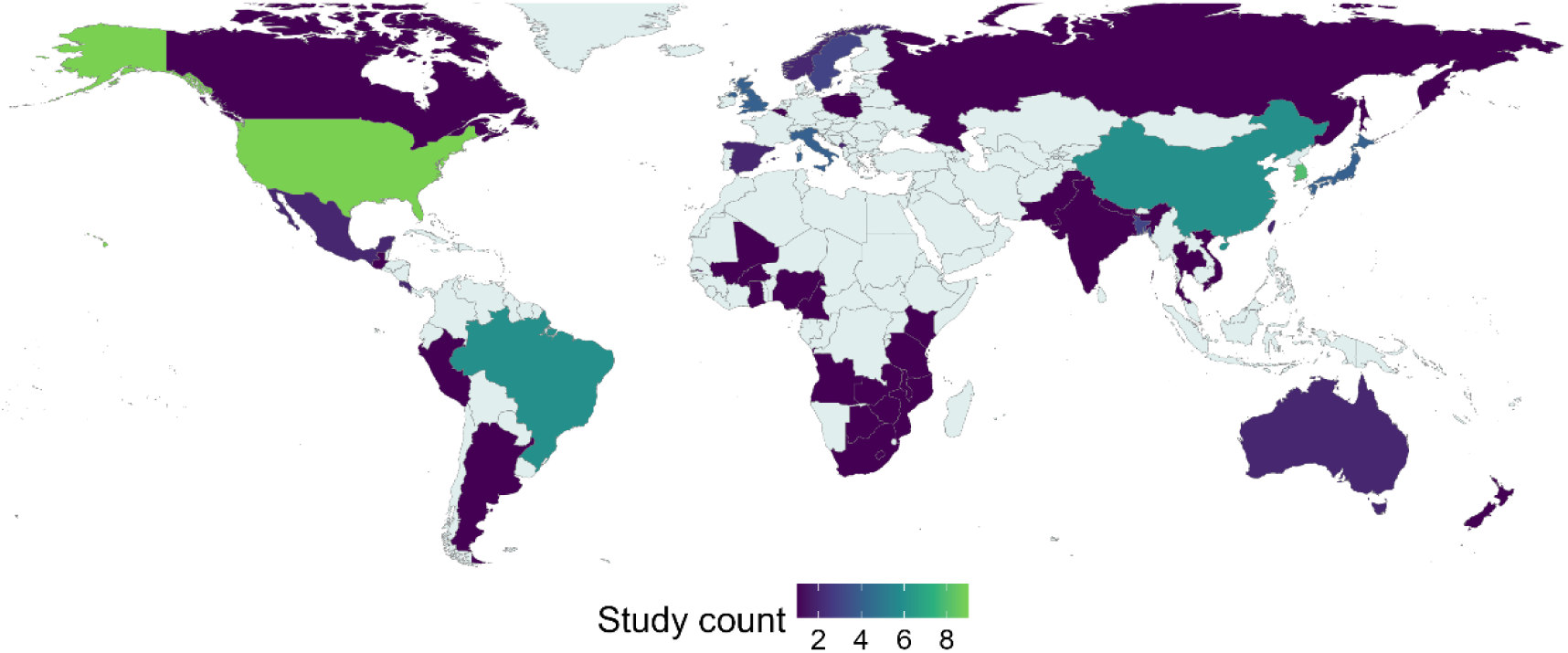
Geographical distribution of included epidemiological and environmental studies.

### 3.2. Outcomes and exposures

We identified the following climate variables as being associated with fluctuations in measurements of NoV burden in human population and in environment settings outside the host: absolute humidity, atmospheric pressure, desiccation, precipitation, radiance, relative humidity, soil moisture (soil runoff), temperature, UV dose and wavelength, vapor pressure, wind speed and directionality. Variables applicable only to environmental settings, i.e., salinity, electrical conductivity, dissolved oxygen or pH, do impact virus survival and behaviour in the environment; but they were excluded as it was not possible to link their variability to incidence of NoV in a human population.

The directionality of associations between measures of NoV incidence risk in human populations and temperature, precipitation or relative humidity were inconsistent across studies of different regions and studies of the same region. This is shown in Table 1, for example, the summarised effect of ambient temperature measured with relative risk was non-significant based on 95% CIs, but significant for rate ratio and regression slope. Even if the relationships for relative humidity and precipitation appear to be consistent and significant (Table 1), it should be noted that many studies did not provide a measure of variance, hence were excluded from the meta-analysis but included in narrative comparisons. Some authors introduced a significant source of bias by calculating the effect only for a sub-interval of the climate variable if the association was non-monotonic. In other cases where NoV incidence risk relative to an explanatory variable was calculated, the numerical boundaries for the latter were not provided, limiting the interpretation of the association. Coupled with highly heterogenous definitions for measurements of NoV incidence risk, incidence or burden in human populations and highly variable definitions of climate variables as outlined in Table S1, comparisons between studies and results from the meta-analysis should be interpreted cautiously and limited to general features, such as directionality.

**Table 1.** Mean effect sizes of individual climate variables on metrics for the measurement of NoV incidence risk and 95% confidence intervals.

| Variable \ Risk Metric | dichotomised |  |  | continuous |  |  |
| --- | --- | --- | --- | --- | --- | --- |
|  | hazard ratio | prevalence ratio | relative risk | pairwise correlation | rate ratio | regression slope |
| **ambient temperature** | - | - | 1.01 (0.94, 1.09) [3] | -0.56 [7] | 0.93 (0.92, 0.94) [3] | -0.0197 (-0.0222, -0.0172) [5] |
| **water temperature** | 5.61 (2.81, 11.12) [1] | - | - | - | 0.93 (0.9, 0.96) [1] | - |
| **relative humidity** | - | - | 0.91 (0.85, 0.96) [2] | -0.23 [4] | 0.98 (0.97, 0.99) [2] | - |
| **precipitation** | - | 0.8 (0.67, 0.93) [2] | 0.88 (0.81, 0.95) [2] | -0.28 [3] | 0.91 (0.85, 0.98) [1] | 0.0493 (0.0194, 0.0792) [4] |
| **absolute humidity** | - | - | - | - | - | -0.1558 (-0.1995, -0.1121) [2] |
| **ambient temperature anomaly** | - | - | - | - | - | 0.0123 (-0.0006, 0.0251) [3] |
| **precipitation anomaly** | - | - | - | - | - | -0.0186 (-0.0674, 0.0303) [3] |
| **atmospheric pressure deviation** | - | - | 0.87 (0.81, 0.93) [1] | - | - | - |
| **soil moisture** | - | - | 1.12 (1.05, 1.19) [1] | - | - | - |
| **wind speed (and offshore versus onshore)** | - | - | 1.13 (1.08, 1.18) [1] | - | - | - |

The numerical format of the measurements is “mean effect (lower 95% confidence interval, upper 95% confidence interval) [number of measurements]” and the results are representative of human populations, but not of NoV measurements outside the host. For temperature, the risk measurements were for a higher temperature relative to a lower temperature, except for the hazard ratio, where a lower temperature group (<4 °C) was compared to a higher temperature group (>4 °C). The units were degrees Celsius (°C) for temperature, % for relative humidity, g of H<sub>2</sub>O per cubic meter or kg of water vapor per kg of dry air for absolute humidity, and mbar for atmospheric pressure deviation. The table includes studies assessed as “Good” and “Adequate” for significance levels marked as “significant” and “not tested”, but excluding “Poor” studies and “insignificant” relationships.

### 3.3. Temperature

With an increase in temperature, generally, there was a decrease in risk of NoV cases, outbreaks or hospitalisations (Figure 4) (Kim & Kim, 2021; Chalapa et al., 2023; Kim et al., 2016; Sung et al., 2022; Kim et al., 2015; Joung, Jang & Kim, 2022; Lopman et al., 2009; Wang, Goggins & Chan, 2018; Ouedraogo et al., 2017; Greer, Drews & Fisman, 2009; Beck-Friis et al., 2023; Mun, 2020; Yang et al., 2022; Wang et al., 2024; Ondrikova et al., 2023; Kim et al., 2021). From laboratory studies, the negative association was explained by quicker reduction rates in the number of virus particles on various fomite types and in liquid mediums at higher temperatures (Figure S1-S2). Similarly, temperature did negatively impact risk of NoV contamination in various types of food products, e.g., oysters, in the laboratory (Figure S3) and was negatively associated with NoV risk in coastal oyster populations and ground, marine and waste waters (Campos et al., 2017; Dienus et al., 2016; Félix et al., 2010; Ilic et al., 2017; Lowther et al., 2012; Miao et al., 2018, 2022; Mohammed, Hameed & Seidu, 2017; Nagarajan et al., 2021; Pérez-Sautu et al., 2012; Rusiñol et al., 2015; Suffredini et al., 2012; Takahashi et al., 2016; Wang & Deng, 2016). In addition to slower decay rates, lower temperatures result in reduced shellfish metabolic activity which likely increases virus bioaccumulation and decrease its clearance rate, facilitating NoV food chain transmission (Table S3; Table S4). However, studies on risk of NoV incidence in human populations hypothesised an additional interpretation of the negative association to be temperature impacting host immunity and population behaviour. Although seasonal variation of temperature does alter human behaviour and social contact patterns, and likely transmission dynamics of NoV, those studies did not provide empirical evidence to confirm this interpretation (Hohm et al., 2024).

**Figure 2.**
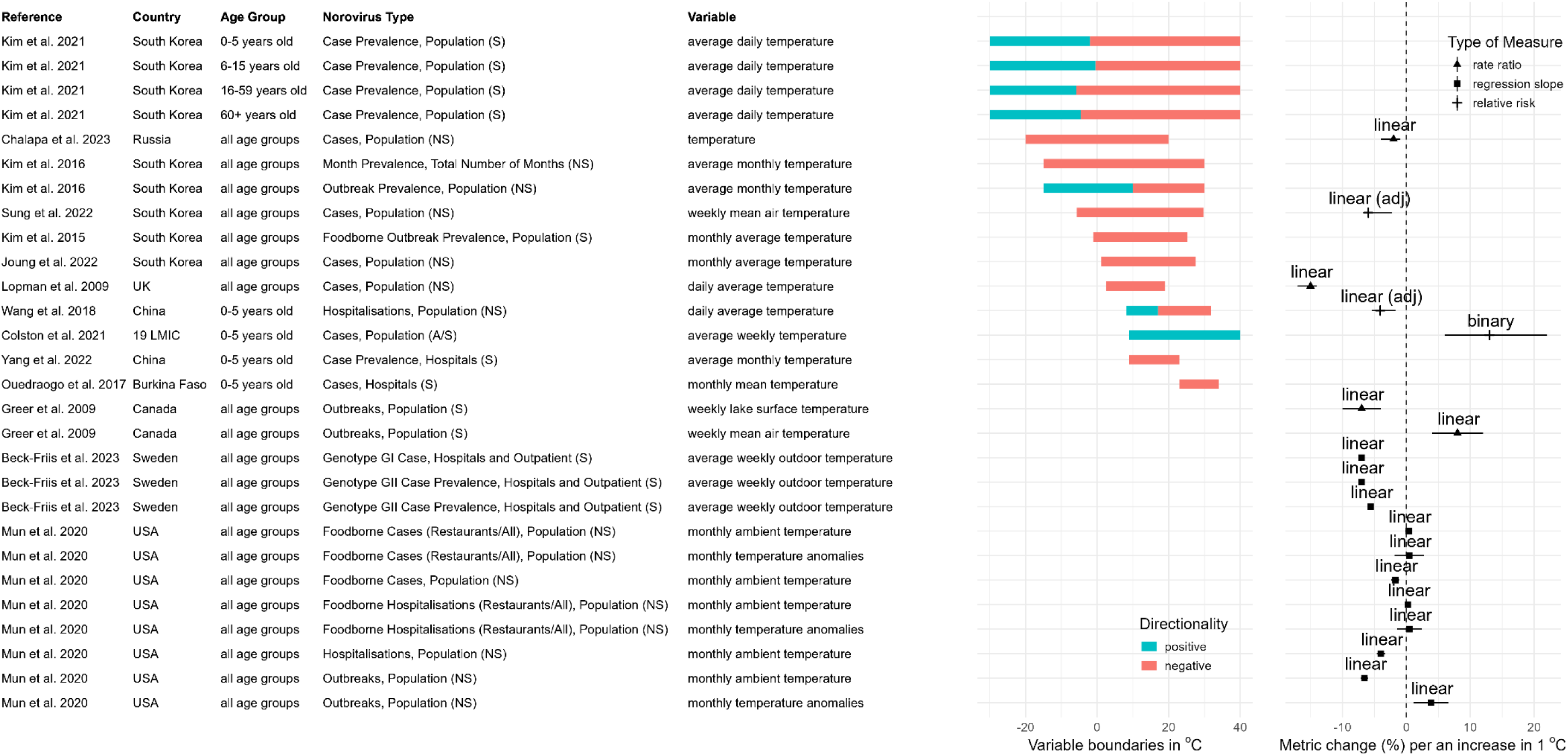
Association of temperature and NoV incidence in human populations. The outcome definition recorded under “Norovirus Type” varied between studies, where RT-qPCR confirmation of a NoV infection was performed on samples from individuals with gastrointestinal symptoms (S), asymptomatic individuals (A), or in the community composed of both asymptomatic and symptomatic individuals (A/S). Some studies used data collected from national surveillance systems and did not specify explicitly their case definition (NS), although they likely reflected mostly patients with gastrointestinal symptoms.

In contrast, in one of the studies by Colston et al. (2022), NoV case datasets were collated from 15 studies across 19 low or middle income countries (LMICs) with variable types of climate, e.g., tropical, subtropical or temperate. They found a positive association between average weekly temperatures and probability of NoV infection (Colston et al., 2022). Contradictorily, two other studies in countries with a tropical climate, Nigeria and Cameroon, found no association (Ayukekbong et al., 2014; Arowolo et al., 2019). The positive association or the lack of any relationship as opposed to a negative association reinforces the possibility that temperature-dependent NoV survivability outside the host is not the sole mechanistic interpretation. The association could originate from other drivers of NoV seasonality, such as changes in population behaviour, including as a response to extreme weather events. For example, two studies demonstrated that anomalies in temperature correlated with a higher number of NoV cases (Mun, 2020; Yin et al., 2024).

### 3.4. Irradiance

Two studies by the same authors found radiance to be a significant predictor of oyster-associated NoV outbreaks in a human population, whereas five other studies showed the association to be insignificant (Shamkhali Chenar & Deng, 2017a, 2017b; Kim & Kim, 2021; Chiu et al., 2022; Chalapa et al., 2023; Colston et al., 2022; Kim et al., 2016). For those two studies, sensitivity analysis indicated that radiance as a predictor is less important than temperature, with a relative contribution of 11-23.8% versus 30-37.2% (Shamkhali Chenar & Deng, 2017a, 2017b). Mechanistically, natural irradiance does accelerate the reduction rate of infectious NoV particles (Figure S6) confirmed by mesocosm studies on effluent wastewater (Park et al., 2021; Ahmed et al., 2024). However, the lack of consensus on its importance with the other studies in larger human populations indicates that (1) the impact of radiance on NoV decay is insufficient to drive the larger seasonality of NoV and that (2) radiance is not a significant predictor of NoV incidence in human populations except in areas proximal to coastal waters.

### 3.5. Precipitation

The direction of the relationship between measures of precipitation and NoV risk of incidence in human populations was both positive and negative with no consensus between studies being able to be deduced (Figure 5) (Colston et al., 2022; Wang, Goggins & Chan, 2018; Bruggink & Marshall, 2010; Joung, Jang & Kim, 2022; Grembi et al., 2024; Greer, Drews & Fisman, 2009; Mun, 2020; Wang et al., 2024). The direct impact of precipitation on NoV incidence will vary significantly depending on the combination of characteristics of the local settlement, such as population density, volume capacity of wastewater treatment plants, proximity to water sources, or even soil properties. When a positive relationship between increased incidence of NoV in the population and higher levels of precipitation was observed, a likely reason was due to contamination of water reservoirs with NoV via soil runoff and combined sewage overflow events (Table S3). For example, Colston et al. (2022) found that oversaturation of soil with moisture from precipitation has also been positively correlated with infection risk of NoV in 19 LMICs.

**Figure 3.**
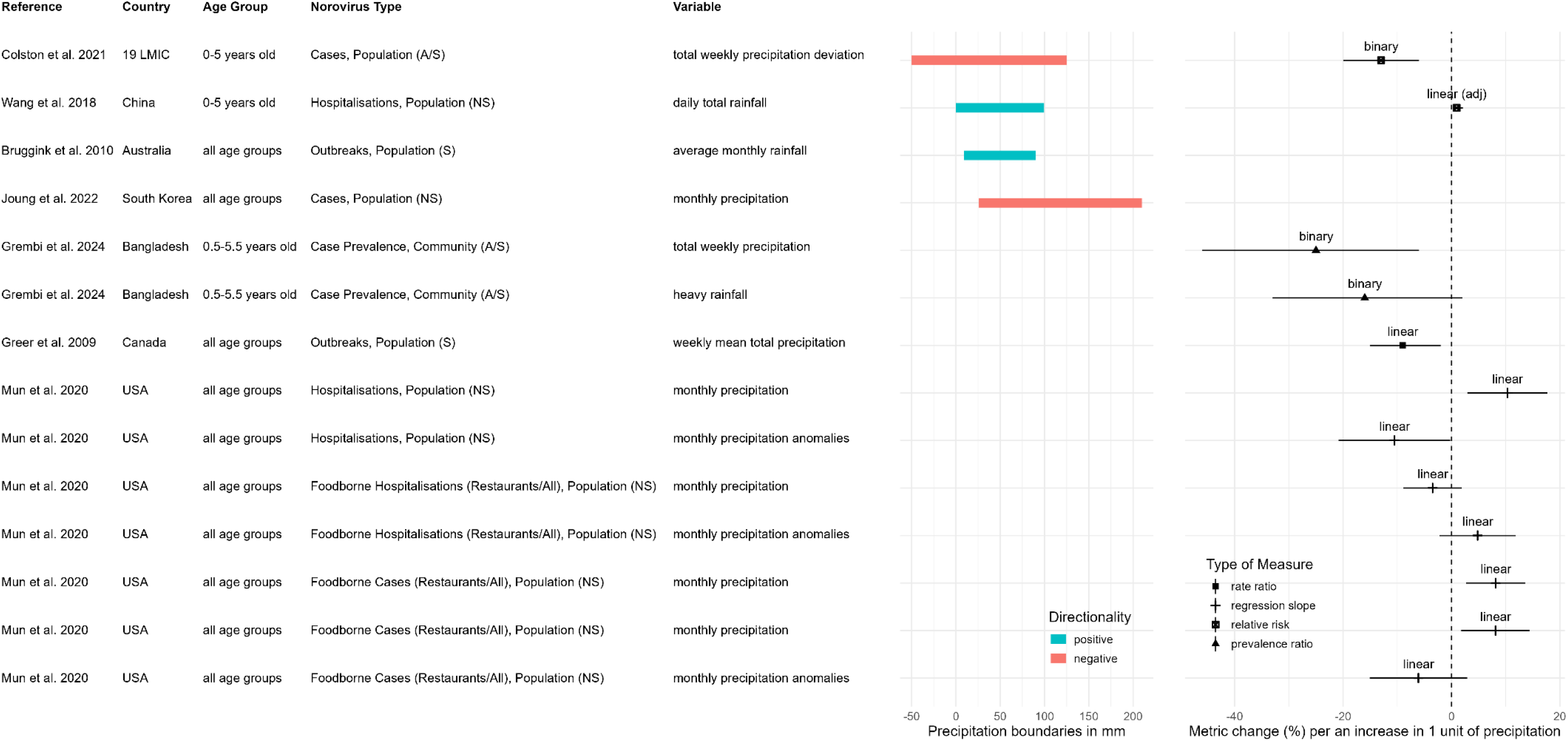
Association of precipitation and NoV incidence in human populations. The outcome definition recorded under “Norovirus Type” varied between studies, where RT-qPCR confirmation of a NoV infection was performed on samples from individuals with gastrointestinal symptoms (S), asymptomatic individuals (A), or in the community composed of both asymptomatic and symptomatic individuals (A/S). Some studies used data collected from national surveillance systems and did not specify explicitly their case definition (NS), although they likely reflected mostly patients with gastrointestinal symptoms.

The direction of the association between precipitation and NoV concentration or outbreak risk in oyster populations were more consistent across environmental studies than when compared to the association of precipitation and NoV incidence in human populations (Figure 5). Among five studies, only one provided a quantitative measure of risk of NoV outbreaks in oysters (Wang & Deng, 2016). Wang et al. (2016) showed that with a rise of 40% in total precipitation levels, the risk of an outbreak increased by 12%. In total, four studies reported a positive association, and one reported a negative relationship (Campos et al., 2017; Nguyen et al., 2018; Tryland et al., 2014; Wang & Deng, 2016; Grković et al., 2017).

The key difference behind the discrepancy in directionality is the assumed mechanism of influence as informed by studies on NoV contamination of environmental reservoirs. In the case of a positive relationship, more precipitation results in the inefficient treatment of sewage followed by the contamination of environmental reservoirs via overflow events and sediment resuspension; whereas for a negative association, more precipitation results in more dilution of the sewage component (de Deus et al., 2019; Graham, Anderson & Boehm, 2021; Hata et al., 2014; Katayama et al., 2004; Kim et al., 2013; Rijal et al., 2009; Rodríguez et al., 2012; Wang et al., 2020; Updyke et al., 2015; Poma et al., 2012; Fumian et al., 2018; Cioffi et al., 2021; Carducci & Verani, 2013). In both cases, trends can also be reflective of seasonal changes of NoV prevalence in the community, such as when driven by seasonal fluctuations in the number of tourists (González-Fernández et al., 2023, 2021; Vieira et al., 2016).

The capacity and quality of wastewater treatment vary significantly between regions, hence, regions with poor processes put in place for the decontamination of household waste will discharge more untreated sewage directly in the environment. In this case, the environmental signal of NoV will be highest in weather with no precipitation and will decrease proportionally with an increase in the amount of precipitation, resulting in fewer NoV particles detected in oyster populations. In comparison, in regions with better sewage treatment infrastructure, discharge of untreated wastewater will take place after the total amount of water arriving at the treatment plant exceeds its processing capacity, usually due to higher amounts of rainfall. Hence, contamination of oysters with untreated sewage and NoV particles is more likely during weather with high precipitation than no precipitation.

These mechanisms of influence could be responsible for the association between precipitation and contamination with NoV of other food supply chains, waters used in irrigation, and recreational groundwaters (de Deus et al., 2019; Hata et al., 2014; Kim et al., 2013; Rijal et al., 2009; Rodríguez et al., 2012; Wang et al., 2020; Rosiles-González et al., 2019). For example, in a study, it was found that 34.38% (22/64) of artisanal cheese produces were positive for NoV in the rainy season versus 17.65% (3/17) in the dry season (Silva et al., 2021). However, no other studies assessed NoV contamination directly in or on fresh produce in the environment relative to precipitation levels. Whereas, if contamination does occur, natural conditions are not sufficient to rapidly reduce the risk of an infection with NoV in fresh produce, requiring >1 day for a 90% number reduction in infectious NoV particles for the 1 °C to 30 °C temperature groups as described in laboratory studies (Figure S3). Even coupling high temperatures (30 °C) to sunlight required a mean time of 55.19 and 11.89 hours for a 90% number reduction of NoV particles on the surface of strawberries and lettuce. In contrast, similar to temperature, some studies in tropical South-East Asia countries report no changes in NoV levels in surface waters or wastewater relative to changes in precipitation (Hoque et al., 2019; Goh et al., 2019).

Besides contamination, one study found that higher precipitation resulted in higher concentration of detected NoV by reducing inhibition of reverse transcription (RT-) real-time polymerase chain reaction (qPCR), highlighting the necessity to account for variability in uncertainty of assays used in environmental surveillance of NoV (Inoue et al., 2020).

### 3.6. Air humidity, vapor and atmospheric pressure

With an increase in relative or absolute humidity a decrease in incidence of NoV was noted (Figure 6) (Beck-Friis et al., 2023; Colston et al., 2022; Chalapa et al., 2023; Joung, Jang & Kim, 2022; Kim et al., 2015; Wang, Goggins & Chan, 2018; Kim et al., 2016; Lopman et al., 2009). Only one study found a positive relationship, and that was conducted in Burkina Faso, which has a dry tropical climate (Ouedraogo et al., 2017). The negative relationship corresponds to the directionality found in laboratory studies where higher relative or absolute humidities resulted in lower NoV survival outside the host (Figure S7). However, this is contradictory to marginally quicker rates of reduction in the number of infectious NoV units in a desiccated medium compared to a control liquid medium (Supplementary Materials 1, “6. Desiccation”). The negative association is also contradictory to the results by Stobnicka-Kupiec et al. (2022). There, relatively higher concentrations of NoV were found in the air at wastewater treatment plants across Poland at higher relative humidities and lower air temperatures, which was attributed to an increased capacity for virus aerosolisation (Stobnicka-Kupiec et al., 2022). As higher relative humidities are associated with higher levels of NoV in aerosols but quicker NoV decay rates, it is possible that (1) an increase in capacity of NoV for aerosolisation does not compensate for quicker reduction in the number of NoV infectious particles, (2) and similar to temperature, air humidity may be an indicator of other drivers of NoV seasonality. When modelling NoV risk of infection via environmental settings, accounting for the impact of environmental conditions not only on the reduction rates of infectious viruses but also on the dynamics of aerosols may provide more accurate estimates of NoV survivability outside the host.

**Figure 4.**
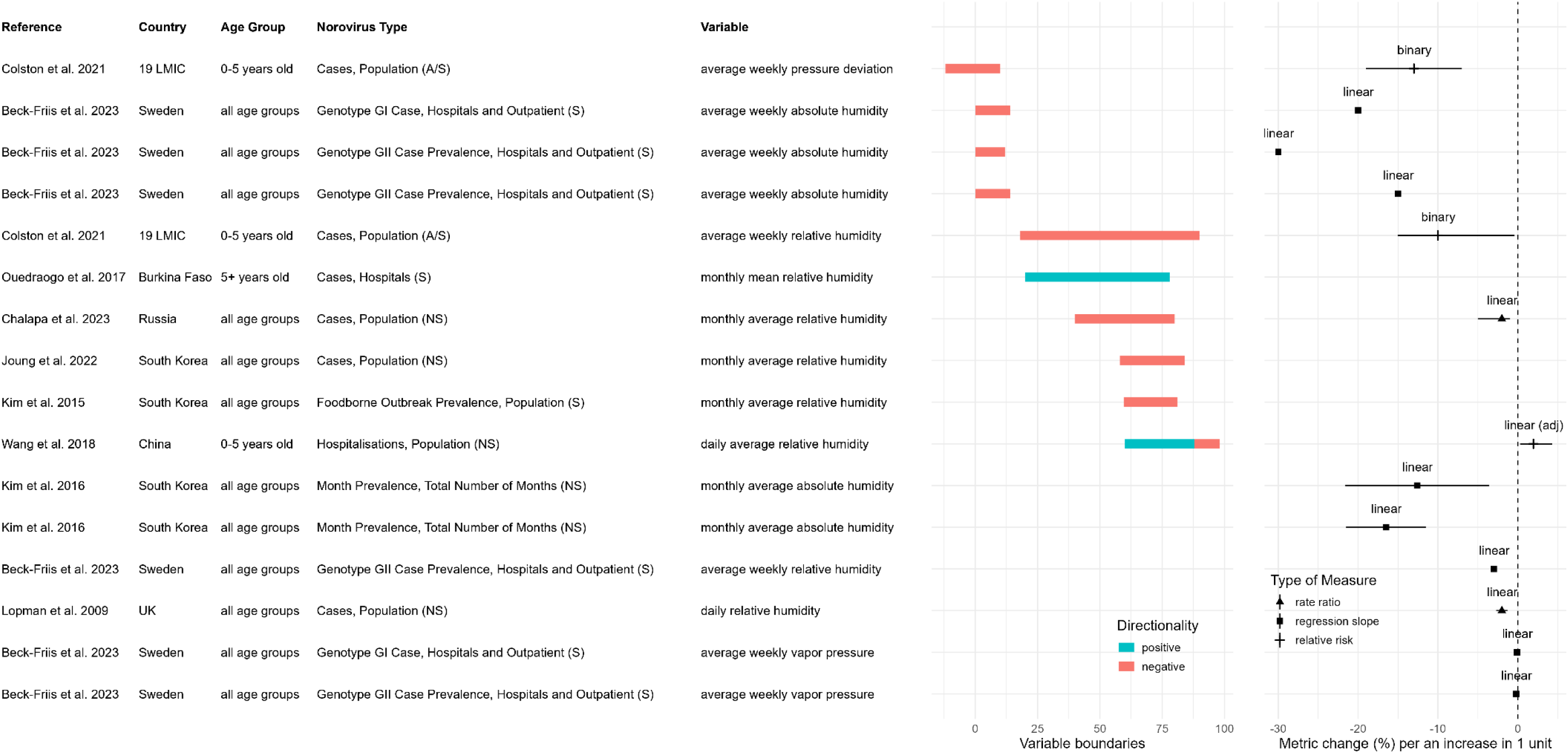
Association of standardised atmospheric pressure, vapor pressure, absolute humidity or relative humidity and NoV incidence in human populations. The outcome definition recorded under “Norovirus Type” varied between studies, where RT-qPCR confirmation of a NoV infection was performed on samples from individuals with gastrointestinal symptoms (S), asymptomatic individuals (A), or in the community composed of both asymptomatic and symptomatic individuals (A/S). Some studies used data collected from national surveillance systems and did not specify explicitly their case definition (NS), although they likely reflected mostly patients with gastrointestinal symptoms. The units are % for relative humidity, g of H_2_O per cubic meter or kg of water vapor per kg of dry air for absolute humidity, mbar for atmospheric pressure deviation, and Pa for vapour pressure.

### 3.6. Atmospheric and vapour pressure

Similarly to air humidity, vapor pressure and deviation of atmospheric pressure also have had a negative association with NoV infection risk (Figure 6) (Colston et al., 2022; Beck-Friis et al., 2023). They have generally been interpreted as proxy indicators for precipitation and absolute or relative humidity; however, that is unlikely to be the case as most regression models adjusted for those as covariates. Hence, to capture the association between climate and NoV incidence risk and decrease the error of regression models, the inclusion of vapor and atmospheric pressure may be necessary, despite of the mechanisms behind these relationship remaining unclear.

### 3.7. Wind

Four studies found wind to be a significant predictor of NoV outbreaks and cases with contradictory directionalities, whereas only one study provided a measure of effect alongside 95% CI (Table 1) (de Almeida et al., 2024; Colston et al., 2022; Shamkhali Chenar & Deng, 2017b; Chenar & Deng, 2018). Relative to other climate factors, the importance of wind was 6.3-9% in predicting oyster associated NoV outbreaks in humans (Shamkhali Chenar & Deng, 2017b; Chenar & Deng, 2018). The main mechanistic interpretation was speculated that wind speed and directionality facilitate the spread of environmental sources contaminated with NoV, but no studies elaborated on whether wind influences other drivers of transmission, particularly social contact patterns, such as recreational time spent near the shore, or whether the association is purely correlative and not causative (Table S3).

### 3.8. Sensitivity analysis

Upon the inclusion of articles classified as poor, the directionality of associations between climate variables and measures of NoV burden for epidemiological studies did not change (Table S5) since no additional studies fulfilled the criteria for inclusion in the meta-analysis due to not providing a measure of dispersion of the mean effect, such as CI or variance.

If excluding poor articles and articles that reported insignificant results or did not report or test significance levels, then the strength of associations based on the slope coefficient of linear regressions increased for relative humidity, precipitation, ambient temperature, and ambient temperature anomalies (Table S6).

## 4. Discussion

Studies reported contradictory results on the significance, directionality, or size of effect for temperature, air humidity, radiance, precipitation, vapour pressure, atmospheric pressure, and wind speed and directionality. When an association was present, individual climate variables were likely temporal indicators of NoV incidence seasonality as well as mechanistically influencing specific pathways of transmission. Although transmission routes of NoV are consistent across different populations, their relative importance is geography specific. Environmental burden and subsequent transmission of NoV is subject to sanitation standards, capacity for wastewater decontamination, food safety procedures, contamination of recreational, irrigation or drinking waters, and even cultural aspects, such as regional nutrition profiles where raw seafood, vegetables and fruits have a higher risk to be contaminated with NoV (Omatola et al., 2024). Where climatic conditions influence NoV burden in the environment, variations in those non-climatic factors will impact the strength of the association between climate variables and total NoV incidence. Heterogeneity of associations also largely stemmed from inconsistent definitions of the exposure variable (e.g., average weekly temperature 7-days before date of notification versus average daily temperature on day of notification), the outcome variable (e.g., NoV cases as a proportion of diarrhoea cases versus total NoV hospitalisations) (Table S1), and the employed statistical approach (e.g., different covariates in linear models), making it difficult to compare and summarise the results quantitatively.

Two other systematic reviews found ambient temperature to be a significant predictor of NoV cases with a rate ratio of 0.89 (95% CI: 0.81-0.99) and a non-significant predictor with a regression coefficient of 0.21 (95% CI: -0.21-0.64) (Chua et al., 2022; Ahmed, Lopman & Levy, 2013). In comparison, we found a similar rate ratio with a mean effect of 0.93 (95% CI: 0.92-0.94) but a significant regression coefficient average estimate of -0.0197 (95% CI: -0.0222 --0.0172) (Table 3). However, the summarised effect is not significant if using relative risk (1.01, 95% CI: 0.94-1.09) (Table 3). We also noted differences in the directionality of the association between continental and temperate climates versus tropical and dry climates, which indicate that the sensitivity of NoV incidence risk to temperature can depend on mechanisms other than higher temperatures reducing the survivability of NoV outside the host. For example, in countries with temperate and continental climates, the school season starts approximately in September, and NoV seasonal outbreaks follow on median 2-3 months later, coinciding with the colder period of the year (Kraut et al., 2017). In contrast, many of the tropical, subtropical and dry climates countries included in this systematic review displayed either a positive association or a lack of it, whereas the start of the school season does not necessarily correspond to the colder months there (Colston et al., 2022; Arowolo et al., 2019; Ayukekbong et al., 2014). Therefore, temperature may be able to capture a range of drivers of NoV seasonality, such as social mixing patters, but the directionality and size of the effect will be country and time period specific.

Alongside temperature, variations in natural radiance, and relative or absolute humidities resulted in different rates of reduction in the number of infectious NoV units outside the host. Since transmission by fomites and food (i.e., fresh produce and shellfish) is an important route of infection, modelling of outside-host NoV dynamics should adjust for virus survival based on those factors (Rushton et al., 2019). Importantly, a study reported that absolute humidity, which can be expressed as a function of temperature and relative humidity, is more critical in influencing NoV survival than relative humidity independently (Colas de la Noue et al., 2014). A consideration is that only two temperature conditions were reported there, at 9 °C and 25 °C, whereas other studies assessing NoV survivability based on varying absolute humidities are not currently available. If assessing the exact burden of NoV in the environment and untangling the transmission rate from direct person-to-person contact, more laboratory or mechanistic modelling studies that estimate the cumulative effect of temperature, relative humidity, absolute humidity and radiance on NoV survivability outside the host are essential.

Regarding precipitation, we found the directionality of the effects to be mixed. Associations in the human population were negative across all metrics except regression slope (Table 3), whereas four out of five environmental studies reported a positive association. In contrast, other systematic reviews found a positive association between more precipitation or the wettest months in the year and higher NoV incidence, likely, consistent with the reported environmental microbiological contamination via combined sewage overflow events (Table S2) (Ahmed, Lopman & Levy, 2013; Mavrouli et al., 2022). Another variable impacting NoV dissemination in the environment is wind speed, which may cause turbulence of environmental waters, thereby resuspending NoV-containing sediment, and may further facilitate the dispersion of wastewater effluent (Chen, 2025). For example, onshore winds have been speculated to drive currents towards the shore, especially affecting oyster farms in the intertidal zone. Regions where wastewater discharge points are adjacent to drinking water sources, waters used for the irrigation of fresh produce, recreational waters, or natural shellfish populations may benefit from using precipitation levels and wind speed and directionality as an advance risk warning of waterborne and foodborne NoV infections (Clough et al., 2026).

Previous reviews explored the potential impact of climate change on NoV transmission by interpreting evidence on the mechanistic influence of individual climate variables (Chen, 2025; Rohayem, 2009). If developing an algorithm projective of NoV incidence risk in a population with climate variables as predictors, reassessing the statistical and mechanistical associations for that specific region is essential to capture the geographical and temporal variability in the relative importance of various pathways of transmission. However, using the effect size from specific regions to project long-term global scenarios of climate change may be an inappropriate application if neglecting variations in the importance of individual routes of environmental NoV transmission.

Lastly, while human NoV could not be previously described in survival assays due to the impossibility to culture it in a cell line (Rzeżutka & Cook, 2004), we found several recent studies that employed infectivity assays directly on human NoV using zebrafish or human intestinal enteroid systems. Considering the availability of these techniques and important differences in the outside-host dynamics of human NoV as opposed to its surrogates, future studies should continue investigating human NoV decay rates by employing a virus culturing step. For environmental surveillance, such as in wastewater-based epidemiology, quantifying NoV with RT-qPCR may still remain the method of choice. RT-qPCR has a higher capacity for high throughput screening and allows to capture more genetic material as opposed to only infectious virus particles, in which case, uncertainty in the environmental signal of NoV may be informed by using decay rates deducted via molecular assays (Walker et al., 2024).

## 5. Conclusion

The directionality and effect size of the associations between individual climate variables and measures of NoV burden in human populations were inconsistent between studies. Variability likely reflected geographical differences of non-climate drivers of NoV transmission, such as differences in seasonality of population behaviour (e.g., the month for the start of the school term, periods of tourism), relative importance of various modes of transmission (e.g., via contaminated seafood), sanitation standards, or wastewater treatment infrastructure. In contrast, NoV survival and dissemination outside the host was highly dependent on climate variables. When modelling burden and transmission of NoV in the environment, the joint effect of temperature, air humidity, irradiance, precipitation and wind speed and direction should be accounted for. However, the extent of their influence on NoV total incidence will be subject to the relative importance of virus transmission via environmental routes versus direct human-to-human contact.

## Data Availability

All data produced in the present work are contained in the manuscript. The data and code analysed in this manuscript are available from the following GitHub repository https://github.com/mrc-ide/climate_norovirus_litreview.

https://www.crd.york.ac.uk/PROSPERO/view/CRD42024628722

## 7. Open Research

The data and code analysed in this manuscript are available from the following GitHub repository https://github.com/mrc-ide/climate_norovirus_litreview.

