## Supplementary Materials 1 for "Associations and mechanisms of influence between climate variables and norovirus seasonal incidence: a systematic review and meta-analysis"

### 1. Infectivity and molecular assays

For the detection and quantification of norovirus (NoV) or its surrogates outside the host in environmental or laboratory studies, we categorised the methods into two, namely infectivity and molecular assays.

In infectivity assays, experiments initially cultured NoV or a surrogate from a sample in a cell line, which only then followed with a measurement of concentration of viruses that are infectious. For example, in some studies, human NoV was proliferated first in a human intestinal enteroid assay or a zebra fish assay, which then followed by quantification of genetic material via reverse transcription (RT-) real-time polymerase chain reaction (qPCR) (Kennedy et al., 2023; Rexin, Rachmadi & Hewitt, 2024; Shaffer et al., 2023; Tan, Gong & Li, 2023). For NoV surrogates, culturing and measurements involved other types of cell lines, such as *E. coli,* feline kidney cells, or murine macrophage lines with measurements of virus concentrations being typically in PFU/ml or 50% tissue culture infectious dose (TCID50). Therefore, the D-value deducted from infectivity assays represents the reduction rate in the number of infectious particles for that particular type of virus. By comparing D-values for NoV or its surrogates at variable climate-like conditions, we are able to understand how important specific variables are in affecting the survivability of the virus and the rate at which the number of infectious units is reduced.

In contrast, molecular assays were categorised as experiments where the detection and quantification of viruses did not involve culturing in a cell line and was performed directly via qPCR or RT-qPCR with or without enzymatic pre-treatment. A key difference of molecular assays from infectivity assays is that they detect and quantify virus particles that are potentially non-infectious and likely overestimate the survivability of the NoV model outside the host due to amplifying genetic material that is not necessarily encapsulated within a virion.

Studies may have addressed this source of bias in molecular assays by coupling RT-qPCR with enzymatic pre-treatment, where free floating genetic material is first degraded enzymatically, followed by the detection of genome only encapsulated and protected by a capsid. In one such case, Rönnqvist et al. (2014) showed that the D-value reduced more than twice from 150 to 60 mJ/cm2 for murine NoV (MNV) and from 300 to 120 mJ/cm2 for NoV genotype GII.4. However, even then, molecular assays are likely to overestimate reduction rates, because when the authors used a plaque assay, which cultures exclusively infectious viruses, the D-value for MNV reduced to 22.67 mJ/cm2 (Rönnqvist et al., 2014). Therefore, D-values deducted via infectivity assays, where NoV models are first cultured in a cell line, represent more accurately the true rate of reduction in the number of infectious viruses.

In recent years, studies were able to culture directly human NoV by using zebrafish and human intestinal enteroid systems. The advantage of these new techniques is the ability to study directly dynamics of infectious human NoV outside the host rather than extrapolate data for it based on surrogates. The rate of reduction in number of infectious particles at similar temperature values were 3.6 to 1490.0 slower for NoV surrogates than for human NoV. This indicates that decay rates of surrogates are not an accurate estimate of human NoV decay in environmental settings. However, for disinfection protocols, assuming higher survivability of human NoV in the environment will decrease the chance of inefficient biohazard decontamination.

### 2. Calculation of D-value

Some studies did not use D-values as a metric of quantifying NoV survivability and rate of reduction outside the human host. Instead, the results might have been presented using alternative metrics of virus concentration or reduction in concentration, e.g., log10 plaque forming units per millilitre (PFU/ml), log10 gene copies per litre (gc/l), log10 reduction, or k-coefficient (slope) of a log10 reduction linear regression trendline, relative to the time since the start of the experiment. In those cases, the data was extracted as given by the study and converted to a D-value as described in the following paragraph.

Let the virus concentration in the control sample or in the sample at the time point 0 be $N_0$ and measurements of virus concentration at subsequent time points be $N_i$, where $i \in \{1, 2, 3, \dots\}$. The log10 reduction in the measurement of virus amount at time points $i$ is then given by $\log_{10}(N_i/N_0)$. After fitting a simple linear regression to the dataset composed of time datapoints (predictor) required to achieve the corresponding log10 reduction in virus amount (predicted variable), we can estimate the $slope$ of the linear regression. The corresponding time required to achieve a 90% reduction in virus concentration, or D-value, is then given by $1/slope$, and the 95%

CIs are given by $1/(slope \pm 1.96 * \hat{\sigma}_s)$, where $\hat{\sigma}_s$ is standard error of the slope mean estimate. Negative D-values or D-values larger than 365 days were excluded from figures as they indicated minimal to no reduction in NoV amount, and because they were outside the time boundaries of all experimental settings. Where applicable, the exposure variables and their effect were also converted to a common type of units, such as Fahrenheit to Celsius.

### 3. Temperature in laboratory studies

Higher temperatures resulted in quicker decay rates with significant variations in D-values between different types of NoV and fomites (Figure S1) (Lee, Zoh & Ko, 2008; Doultree et al., 1999; Colas de la Noue et al., 2014; Lamhoujeb et al., 2009; Liu et al., 2012; Kim et al., 2012; Samandoulgou et al., 2015; Wollants et al., 2004; Abou-Hamad et al., 2023). MNV and FCV had reduction rates closely aligned to each other and much lower than those for MS2.

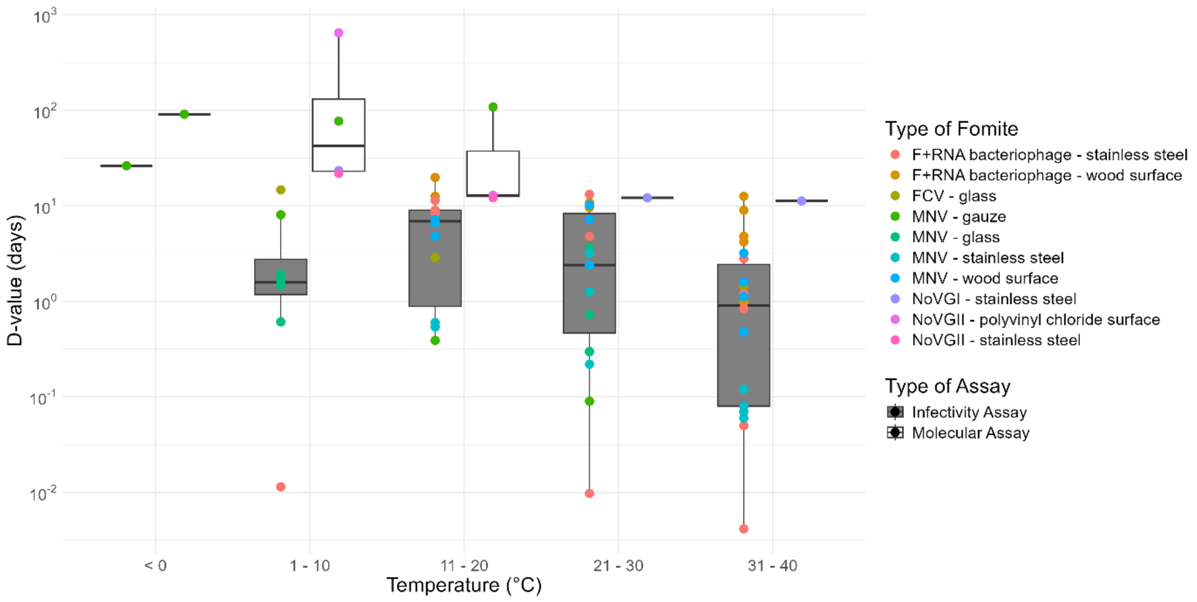


*Figure S1. D-values of NoV and NoV surrogates measured at variable temperatures on fomites.*

In liquid mediums, reduction in the number of infectious particles at comparable temperature ranges were slower than on fomites (Figure S2) and is likely one of the main mechanism responsible for the negative association between temperature and NoV concentration in environmental studies (data not shown) (Allwood et al., 2003; Araud et al., 2016; Bae & Schwab, 2008; Brié et al., 2017; Cannon et al., 2006; Doultree et al., 1999; Duizer et al., 2004; Fu et al., 2023; Hewitt, Rivera-Aban & Greening, 2009; Ibrahim et al., 2019; Kauppinen & Miettinen, 2017; Keller et al., 2010; Kennedy et al., 2023; Kennedy, Lowry & Boehm, 2024; Liu et al., 2012; Park, Bae & Ha, 2015; Rexin, Rachmadi & Hewitt, 2024; Seo et al., 2012; Shaffer et al., 2023; Tiwari et al., 2023;

Tuladhar et al., 2012; Verhaelen et al., 2012; Zhu et al., 2020; Skraber et al., 2009; Dean et al., 2020; Gibson & Schwab, 2011; Ao et al., 2019; Tubatsi & Kebaabetswe, 2022). Another mechanism of temperature impacting NoV levels in environmental water sources is bacteria, such as *S. multivorum*, removing NoV the most optimally at 25 °C (Yu et al., 2024).

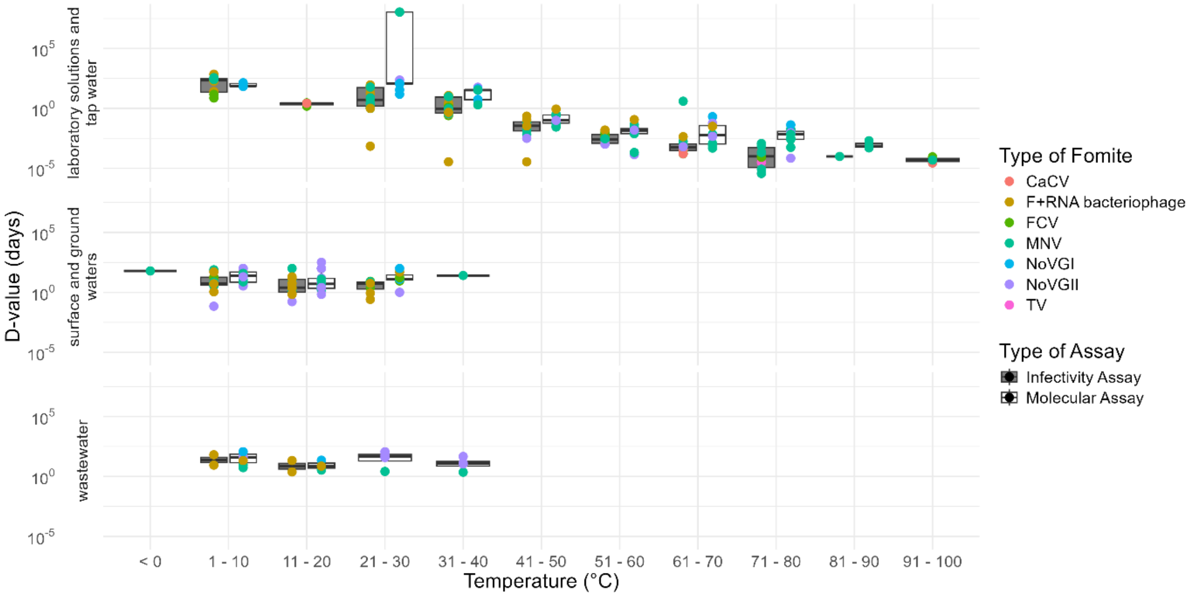


*Figure S2. D-values of NoV and NoV surrogates measured at different temperatures in liquid environments.*

Similarly, temperature had a negative impact on the decay rate of NoV on the surface of vegetables and fruits, and on the contamination levels with NoV in oyster viscera or midgut gland and turkey meat (Figure S3) (Bozkurt, D'Souza & Davidson, 2015; Carratalà et al., 2013; Choi & Kingsley, 2016; Dhulappanavar & Gibson, 2024; El-Senousy et al., 2020; Hewitt, Rivera-Aban & Greening, 2009; Lee et al., 2015; Park, Bae & Ha, 2015; Rupnik et al., 2021; Stoppel et al., 2023; Verhaelen et al., 2012; Younger et al., 2020). In the top panel of Figure S3, we also included laboratory experiments on the impact of temperature on oyster depuration rates. In addition to NoV decay, depuration involves the biological removal of NoV not only via NoV decay but also by oysters physiologically filtering out contaminations. However, for contrast, some studies on NoV risk in mussel and shellfish populations report no correlation between risk of NoV contamination and ambient or water temperature (Bazzardi et al., 2014; Grković et al., 2017; Hernroth et al., 2002).

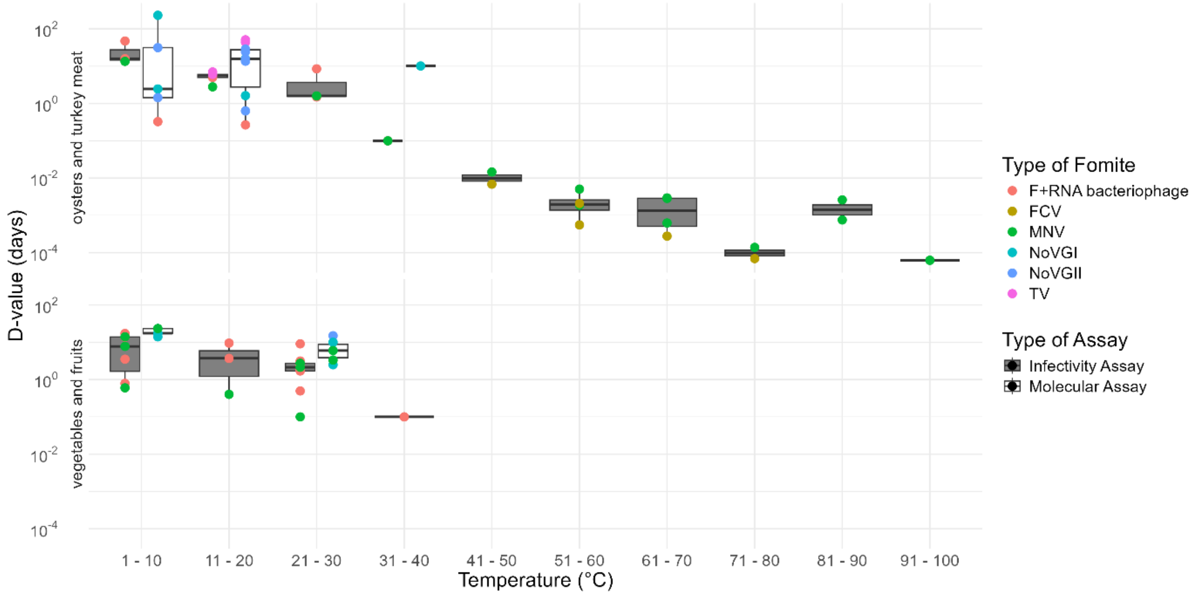


*Figure S3. D-values of NoV and NoV surrogates measured at different temperature conditions in contaminated oysters and turkey meat (top panel), and vegetable and fruits (bottom panel).*

In artificial stool and on artificial diaper material, the D-value decreased exponentially with an increase in temperature based on plaque assay experiments (Cannon et al., 2006; Lee, Zoh & Ko, 2008). At -20 °C, the D-values for diaper material and stool suspension were comparable between RT-qPCR and plaque assay, denoting limited to no decay (Auffret et al., 2019; Baert et al., 2010; Cannon et al., 2006; Lee, Zoh & Ko, 2008). Interestingly, for all temperatures except -20 degrees, decay rates in stool suspensions were higher than on diaper material with values of 232.34, 16.05 and 0.3 days versus 21.94, 1.28 and 0.24 days at 4, 18 and 30 degrees.

No protective effect against reduction of viral infectivity could be observed for stool suspension, or for diaper material if the virus is transported onto it, compared to other types of fomites or buffer solutions. The slope of the temperature-dependent D-value exponential trendlines were lower for gauze and stainless steel and higher for glass and wood fomites, corresponding to relatively slower and, respectively, quicker decay rates for diaper material. Similarly, the decay rate was similar or faster in stool suspension than in buffer solutions. An artificial stool environment did not offer additional protection to FCV from desiccation on stainless steel than on a glass fomite when comparing similar temperature conditions. At 4 °C and 20-22 °C, the D-values were 5.46 and 2.38 days for artificial stool on stainless steel versus 15.16 and 3.9 days on glass.

No laboratory articles described the impact of temperature on aerosol dynamics and NoV survivability. However, the results on fomite mediums can be used to interpret reduction rate of NoV infectious particles in aerosols upon settling on a surface. Some of the studies allowed droplets to dry first before testing the effect of the exposure condition, while other experimental set-ups prevented evaporation before and during the testing. For example, in regions with hot and dry climates, aerosol evaporation will occur very quickly, and included fomite studies that allowed the droplets to dry first will be more suitable in assessing the duration over which infectious particles are preserved in aerosols.

### 4. Irradiance and ultraviolet light exposure in laboratory studies

With an increase in dose of exposure to UV light, the log10 reduction of the NoV model accelerates (Mitchell et al., 2019; Rockey et al., 2020). Despite a limited number of studies, lower UV wavelengths required smaller doses of exposure to achieve a one log10 reduction in the number of infectious virus particles with mean D-values of 8.96, 16.13 and 12.59 mJ/cm2 for 254, 260 and 279 nm (Polen et al., 2024; Corson et al., 2024; Rönnqvist et al., 2014; Park et al., 2015). Wavelengths of lower amplitudes, as measured by nanometres (nm), have higher frequencies and possess higher electromagnetic energies (Vo, Hernandez & Patel, 2025). In turn, lower wavelengths will generally require lower light intensities or light energy required by the emitting device (mJ/cm2) to achieve similar damage on nucleic acids as irradiance with a wavelength of higher amplitudes. It should be noted that radiance at these wavelengths (Figure S4) is filtered out by the Earth's atmosphere, whereas no studies used UV conditions above >279 nm on fomites.

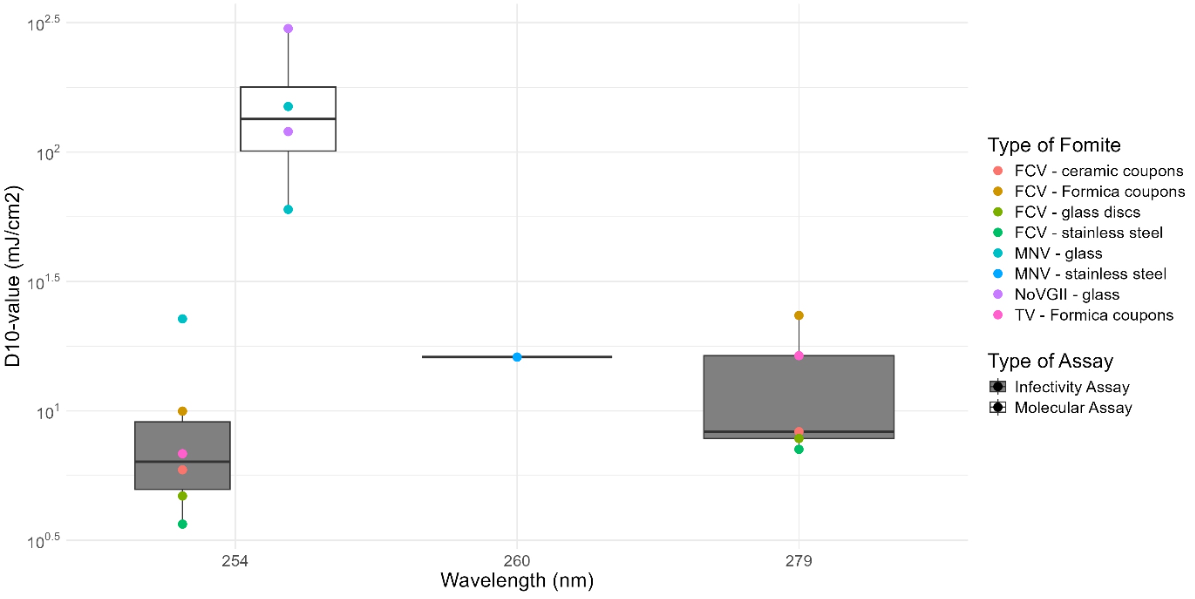


*Figure S4. D10-values or D-values of NoV and NoV surrogates measured at different UV wavelengths on fomites.*

In liquid mediums, lower wavelengths required less exposure time to irradiance to achieve a reduction of one log10 in the number of infectious particles, with mean D-values of 7.32-14.08 mJ/cm2 for 220-280 nm (Figure S5) (Araud et al., 2020; De Roda Husman et al., 2004; Duizer et al., 2004; Lee, Zoh & Ko, 2008; Lee & Ko, 2013; Park, Linden & Sobsey, 2011; Tan, Gong & Li, 2023; Walker et al., 2019; Weng et al., 2018; Yoon, Kim & Cho, 2023; Zhang et al., 2021; Zhu et al., 2020; Samandoulgou, Fliss & Jean, 2015, 2021). In comparison, the wavelengths in the range of UV-B (280-315 nm) and UV-A (315-400 nm) light required significantly more irradiation with D-values of 129.01 and 9747.47 mJ/cm2.

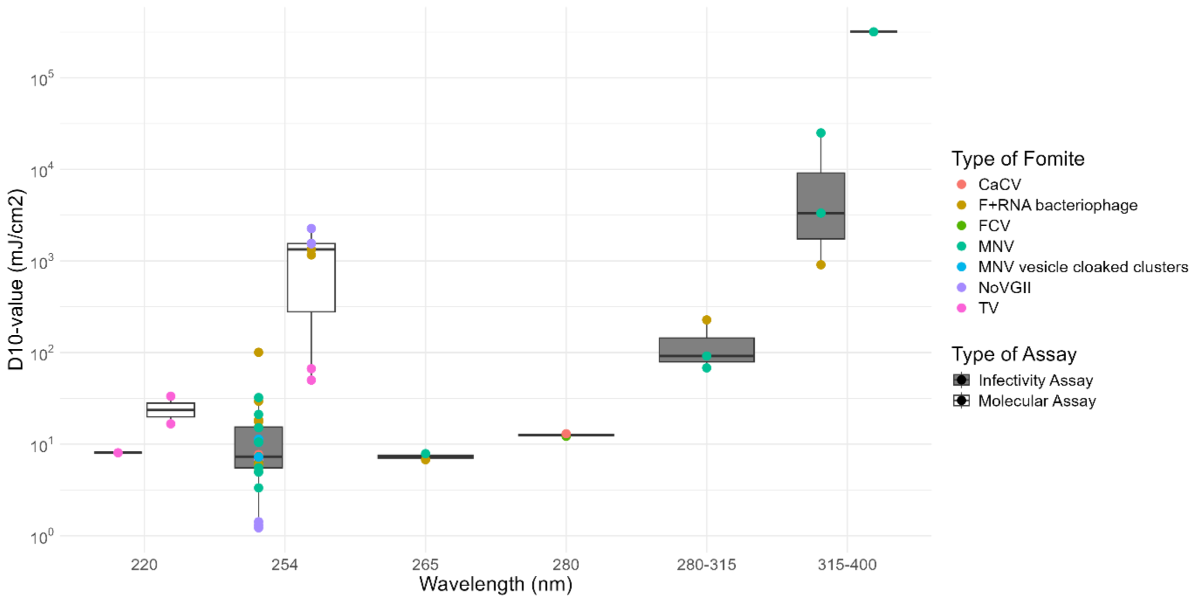


*Figure S5. D10-values or D-values of NoV and NoV surrogates measured at different UV wavelengths in liquid environments.*

Laboratory experiments demonstrated that the impact of natural or natural-like radiance on the reduction of NoV infectivity is significant (Figure S6) (Tiwari et al., 2023; Loeb et al., 2021; Flannery et al., 2013; Elmahdy et al., 2018). Furthermore, one of the studies measured that the total natural radiance required for a one log10 reduction in the number of infectious virus particles corresponds to 20000 mJ/cm2 at 17 degrees Celsius (Flannery et al., 2013). Given that the average daily dose of UV-B and UV-A light can be 1280 mJ/cm2 and, respectively, 42530 mJ/cm2, depending on the region and year (data for Reading, UK, 1998), natural radiance is a significant factor in accelerating the reduction of NoV infectivity in liquid environments as well as on fomites (Kift et al., 2006).

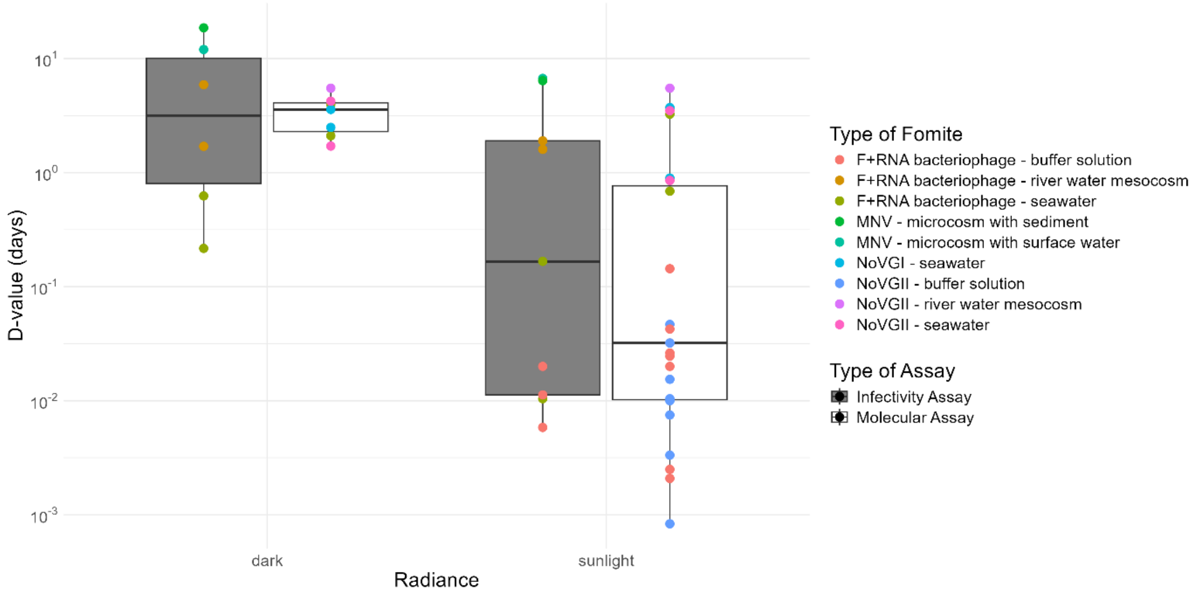


*Figure S61. D-values of NoV and NoV surrogates in liquid environments when exposed to natural irradiation (sunlight) versus control (dark) conditions.*

### 5. Air humidity in laboratory studies

Optimal conditions for preserving a higher number of infectious NoV particles were at a relative humidity of 0-20% followed by 21-40% with a mean D-value of 10.10 and 4.89 days, whereas the highest rate of reduction was at a relative humidity of 61-80% with a mean D-value of 2.91 days (Figure S7) (Buckley et al., 2017; Colas de la Noue et al., 2014; Lamhoujeb et al., 2009; Kim et al., 2012; Girard et al., 2010). Between 81-100%, the reduction rate of infectious particles was marginally slower than at 61-80%.

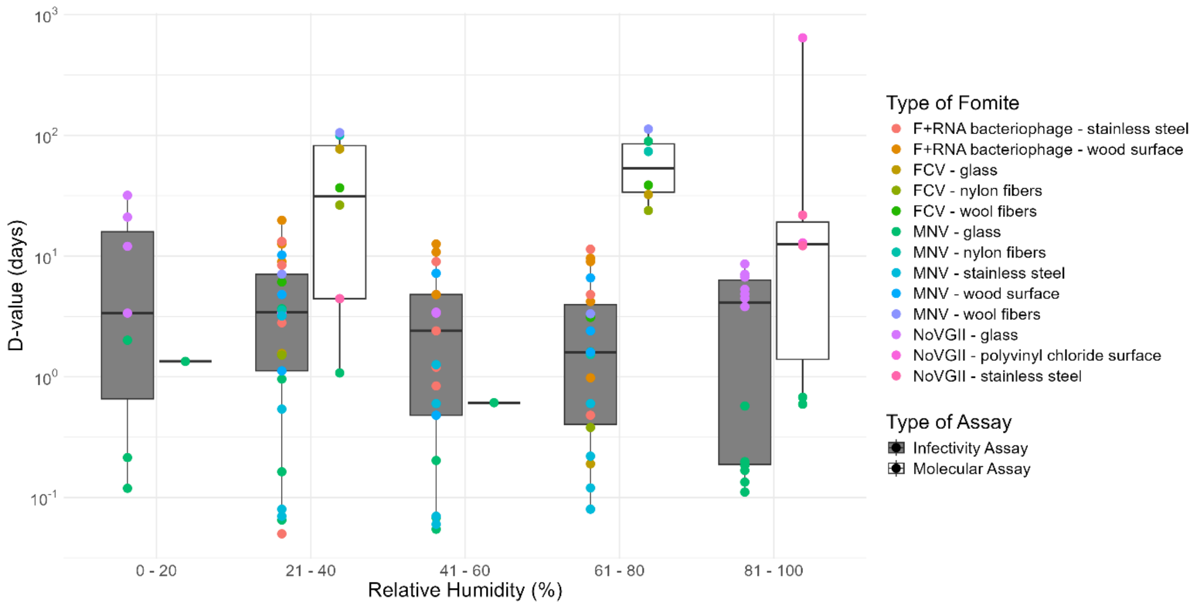


*Figure S7. D-values of NoV and NoV surrogates measured at variable relative humidities on fomites.*

Only one study measured the effect of absolute humidity, which was reported to be more important than relative humidity in affecting the reduction of NoV infectivity on fomites (Colas de la Noue et al., 2014). Colas de la Noue et al. (2014) noted that below 0.007 kg water/kg air were favourable conditions, whereas values about this threshold would result in much lower virus survival.

In agricultural settings, relative humidity may also impact the extent of contamination of fresh produce. Besides decay, air humidity affects plant transpiration and internalisation levels of NoV from its surface. One such study showed that as the relative humidity increased from 70% to 99%, the plant transpiration decreased from 0.032 to 0.0031 g/cm2/h, resulting in a decrease in the prevalence of NoV on its surface from 7/8 to 1/8 replicates (Wei et al., 2011).

### 6. Desiccation

When the liquid evaporates completely, which is more likely to occur at lower air humidities and higher temperatures, the medium becomes desiccated, and the dynamics of NoV survivability may change. However, desiccation appeared to have a minimal effect on influencing the reduction in the number of infectious virus units, ambient temperature being much more significant. For example, the D-values for the desiccated medium were 15.16, 3.90 and 1.21 at 4, 20 and 37 $^{o}$C versus 14.67, 2.86 and 1.24 in liquid suspension (Doultree et al., 1999). In contrast, studies on NoV in artificial stool as a medium showed that the reduction rates in the number of infectious particles for NoV surrogates, MNV and FCV, was generally quicker when dried versus than when in a liquid suspension with D-values between 4.11-5.46 versus 5.66-9.59 at 4 $^{o}$C and 1.5-2.38 versus 2.04-4.24 at 22 $^{o}$C (Cannon et al., 2006).
